# Population Attributable Mortality Associated with Respiratory Viruses in Ontario

**DOI:** 10.64898/2025.12.27.25343084

**Authors:** David N. Fisman, Alicia A. Grima, Natalie J. Wilson, Simran Mann, Ashleigh R. Tuite, Clara Eunyoung Lee

## Abstract

**Background:** Respiratory viruses are major contributors to population mortality, but cause-of-death coding undercounts their impact. Ecological regression models linking viral circulation to mortality fluctuations can address this limitation.

**Aim:** To estimate the population attributable fraction (PAF) of mortality associated with influenza A and B, respiratory syncytial virus (RSV), and SARS-CoV-2 in Ontario, Canada (1993–2025), and to characterize temporal changes in virus-attributable mortality across the pandemic transition.

**Methods:** We analysed monthly all-cause mortality data with laboratory surveillance indicators for influenza A, B, RSV, and SARS-CoV-2. Negative binomial models with secular trends, Fourier seasonal terms, and population offsets were fit for pre-pandemic (January 1993–February 2020) and combined pandemic (March 2020–February 2025) periods. PAFs were derived from counterfactual predictions setting viral coefficients to zero. Sensitivity analyses examined temporal stratification of the pandemic period (Public Health Emergency of International Concern [PHEIC] period: March 2020–April 2023; post-PHEIC: May 2023–February 2025), exclusion of the early pandemic period (March–June 2020), and models without Fourier seasonal adjustment. Wald tests compared coefficients across specifications.

**Results:** Pre-pandemic, influenza A accounted for 1.8% (95% CI 1.4–2.3%) of mortality; influenza B showed no detectable impact. RSV demonstrated inverse associations in seasonally adjusted models but positive associations (PAF 1.9%, 95% CI 1.3–2.4%) without seasonal adjustment. Over the combined pandemic period (March 2020–February 2025; n=60 months), amid elevated baseline mortality (IRR 1.050, P=0.027), SARS-CoV-2 accounted for 6.1% (95% CI 4.2–8.0%) of deaths, approximately 4-fold the pre-pandemic influenza A burden, despite widespread vaccination and antiviral availability. Model-estimated SARS-CoV-2-attributable deaths closely matched reported COVID-19 deaths from Public Health Ontario over the same period. Temporal stratification identified a significant increase in SARS-CoV-2-attributable mortality in the post-PHEIC period (PAF 9.8%, 95% CI 1.1–17.7%; p=0.027), while post-PHEIC influenza A and B attributable fractions did not differ significantly from pre-pandemic baselines. Excluding March-June 2020 yielded a conservative SARS-CoV-2 PAF of 5.7% (95% CI 3.3–8.1%), confirming robustness of primary estimates. Meta-analyses showed substantial heterogeneity for influenza A (I²=92.8%) and RSV (I²=89.1%) across modeling approaches, but minimal heterogeneity for SARS-CoV-2 (I²=5.5%).

**Conclusion:** SARS-CoV-2 was associated with a 3–4-fold higher population mortality burden than seasonal influenza A despite available countermeasures. Post-PHEIC data suggest that the burden of respiratory virus mortality, including for influenza, has not returned to pre-pandemic levels, highlighting the continued importance of respiratory virus prevention strategies. Estimates for influenza A and RSV were sensitive to seasonal adjustment, highlighting the importance of modelling choices when quantifying virus-attributable mortality.

## Introduction

Respiratory viral illnesses impose a substantial burden on individuals and the health care system, often resulting in significant morbidity and mortality. Many of these viruses are now vaccine preventable or can have their severity attenuated through vaccination. This holds true in a Canadian context, where two of the leading causes of death are respiratory illnesses; influenza, which prior to the COVID-19 pandemic, caused approximately 3,500 deaths annually (1), and severe acute respiratory syndrome coronavirus 2 (SARS-CoV-2), which as of 2023 had caused a reported 55,282 deaths in Canada (2, 3). Other respiratory viruses, such as respiratory syncytial virus (RSV), can also cause substantial morbidity and mortality, particularly in vulnerable ages groups (e.g., children and older adults) (4–6).

To prioritize vaccination and other prevention programs against respiratory viruses, it is important to understand the potential for prevention of morbidity and mortality at the population level. One such method of doing so is by analysing the population-attributable fraction (PAF) for a given respiratory virus. PAF quantifies the proportion of cases that could be prevented if the exposure, (in this case, the viral infection), were eliminated, offering a valuable metric for public health planning (7). PAF can be calculated as the difference between the observed number of cases (O) and the expected number of cases if the exposure was removed (E), divided by the observed number of cases (PAF = (O-E)/O) (7). “Shortcut formulae” allow PAF to be estimated if the relative risk among the exposed, and the prevalence of exposure, are known (7, 8).

When mortality is the outcome of interest, researchers often rely on cause-specific mortality data to estimate the impact of respiratory viruses. However, these data are subject to well-known limitations, including misclassification and inaccuracies in death certificate coding (9).As an alternative, regression-based methods offer an indirect means of estimating the PAF (10).These approaches do not require explicit cause-of-death attribution and are agnostic to the magnitude of the exposure effect. By modeling the statistical association between viral circulation and mortality, often using smoothing techniques to detrend and smooth data, such models can estimate the fraction of mortality attributable to a given virus, based on the observed changes in outcomes corresponding to fluctuations in exposure.

In this study, we sought to quantify the population-level mortality burden attributable to key respiratory viruses in Ontario, Canada, using regression-based approaches. Specifically, we estimated the PAF for influenza A and B, and respiratory syncytial virus (RSV) over the period from 1993 to 2025, and for severe acute respiratory syndrome coronavirus 2 (SARS-CoV-2) from 2020 to 2025. By applying statistical models that account for temporal variation in viral activity and mortality, our aim was to provide robust, population-based estimates of virus-attributable mortality to inform future vaccination strategies and respiratory virus prevention policies. We also evaluated how modelling choices, specifically the inclusion or exclusion of seasonal adjustment term, affect estimates of virus-attributable mortality.

## Methods

### Data Sources

We calculated percent positivity for influenza A and B, and RSV in Ontario using laboratory-based surveillance data from FluWatch, Canada’s national surveillance system for influenza and other respiratory viruses, available from 1993 onward (1). For SARS-CoV-2, we used test-adjusted case counts from Ontario’s Case and Contact Management System (CCM) (11) from March 2020 to August 2022 (using methodology described in Fisman et al. (12) and Bosco et al. (13)) and percent positivity from FluWatch from September 2022 onward. Both measures were included in pandemic-period models to account for the surveillance system transition from universal to targeted testing; the counterfactual sets both to zero when estimating SARS-CoV-2 PAF. Mortality data were obtained from Statistics Canada (1991–1993) and the Ontario Deaths Registry (1994–February 2025) (14, 15). Data were aggregated to monthly intervals; monthly aggregation reduces noise in mortality attribution models and aligns with established influenza mortality attribution literature (10). Population denominators were obtained from Statistics Canada (14). Individual-level CCM data are not publicly available; all other data sources are public, and aggregated monthly data used in this analysis are provided in a public repository (see Data Availability).

### Analysis

All analyses were conducted using Stata version 18. To estimate the number of deaths attributable to seasonal respiratory viruses and SARS-CoV-2, we fit regression models to monthly all-cause mortality data from Ontario, incorporating indicators of viral circulation. We modeled death counts using both Poisson and negative binomial regression, including Fast Fourier Transforms (FFT; sine and cosine terms) to account for seasonal periodicity, and linear and quadratic terms to model secular trends. Population denominators were included as a log-offset to estimate mortality rates.

Poisson model fit was assessed using Pearson and deviance goodness-of-fit tests (estat gof in Stata). Evidence of overdispersion (p < 0.001 for both tests) supported the use of negative binomial regression for all further analysis. Given that SARS-CoV-2 did not circulate widely before March 2020, analyses were stratified into pre-pandemic (January 1993–February 2020) and pandemic (March 2020–February 2025) periods. The pandemic period was defined as extending through February 2025, the most recent month for which mortality data were assessed as substantially complete; mortality data for subsequent months showed evidence of reporting lag and were excluded. Secular trend terms were centred within each period to reduce collinearity.

As a sensitivity analysis, we re-estimated all models excluding Fourier terms to evaluate the robustness of viral effect estimates to the inclusion of seasonal periodicity. Seasonal adjustment terms may compete with viral predictors for explanatory power, particularly when viruses exhibit strong seasonal patterns. The reduced models were otherwise identical in structure, retaining trend terms, population offsets, and viral covariates.

To formally assess whether removal of Fourier transforms significantly altered viral effect estimates, we compared regression coefficients between primary and sensitivity models using Wald tests. Seemingly unrelated estimation (suest command in Stata) was used to jointly estimate both models and obtain the variance-covariance matrix needed for hypothesis testing. For each viral covariate, we tested the null hypothesis that coefficients were equal across models. P-values < 0.05 were considered evidence of significant modification by seasonal adjustment.

For both primary and sensitivity models, we estimated the fraction of all-cause mortality attributable to each virus by generating model-based predictions under two scenarios: one with observed viral circulation, and a counterfactual in which the coefficients for each virus (or all viruses) were set to zero. PAF was then calculated as the difference between predicted deaths under observed and counterfactual scenarios, divided by the predicted deaths under observed conditions. Confidence intervals for each PAF estimate were obtained using the punaf command, specifying virus-specific terms to be set to zero in the counterfactual. We validated model-based estimates by comparing them to manually derived estimates from predicted mortality totals.

We conducted three pre-specified sensitivity analyses in addition to the Fourier comparison described above. First, we stratified the pandemic period into the SARS-CoV-2 Public Health Emergency of International Concern (PHEIC) period (March 2020–April 2023) and post-PHEIC period (May 2023–February 2025). Given the smaller sample size of each stratum (n=38 and n=22 months, respectively), these temporal comparisons are presented as exploratory analyses; the combined pandemic model (n=60) is treated as primary. Second, we excluded March–June 2020 from the pandemic period to assess the influence of the early pandemic phase, characterised by health system disruption, incomplete surveillance, and high case-fatality rates prior to vaccination (July 2020–February 2025; n=56 months). Third, we compared model fit using adjusted versus unadjusted SARS-CoV-2 case counts using the Akaike and Bayesian Information Criteria (AIC/BIC), with the model using test-adjusted counts demonstrating better fit (AIC = 5145.2 vs. 5162.1). Adjusted counts were therefore retained for primary analyses.

### Meta-analysis

We conducted random-effects meta-analyses to generate pooled estimates of the population attributable fraction (PAF) for each viral pathogen individually, and for all respiratory viruses combined, stratified by time-period and model specification. For each virus or virus grouping, and for each combination of modeling approach (with or without Fourier terms) and time-period (pre-pandemic vs. intra-pandemic), we extracted PAF estimates and 95% confidence intervals from counterfactual models using the punaf command in Stata. Meta-analyses were performed using the DerSimonian and Laird method with inverse-variance weighting, as implemented via the metan command in Stata. We assessed heterogeneity using Cochran’s Q test and the I² statistic. Stratum-specific estimates were visualized using forest plots.

Significant heterogeneity was observed for estimates associated with all respiratory viruses combined, as well as for influenza A and respiratory syncytial virus (RSV). To explore sources of heterogeneity, we conducted subgroup meta-analyses stratified by inclusion of Fourier terms and by pandemic period, holding the other factor constant. P-values for between-group heterogeneity were calculated for each comparison.

## Ethics and Data Availability

All data used in this study were aggregate-level, with viral positivity data and monthly mortality being publicly available. All data and analysis code are available at https://github.com/fismanda/respiratory-virus-mortality-ontario (archived at https://zenodo.org/records/19035861). Approval for use of aggregate level, test-adjusted SARS-CoV-2 case counts was granted by the University of Toronto Research Ethics Board (Protocol # 41690). Age-specific mortality attribution will be addressed in future work.

## RESULTS

### Model Fit

Analyses were stratified into pre-pandemic (January 1993–February 2020) and pandemic (March 2020–February 2025) periods. Negative binomial regression demonstrated superior fit to Poisson models (p < 0.001 for overdispersion), and the pandemic period itself was associated with elevated all-cause mortality (IRR 1.050, 95% CI 1.005-1.084, P = 0.027). Mortality and virological surveillance data are shown in **Figure 1**.

**Figure 1.**
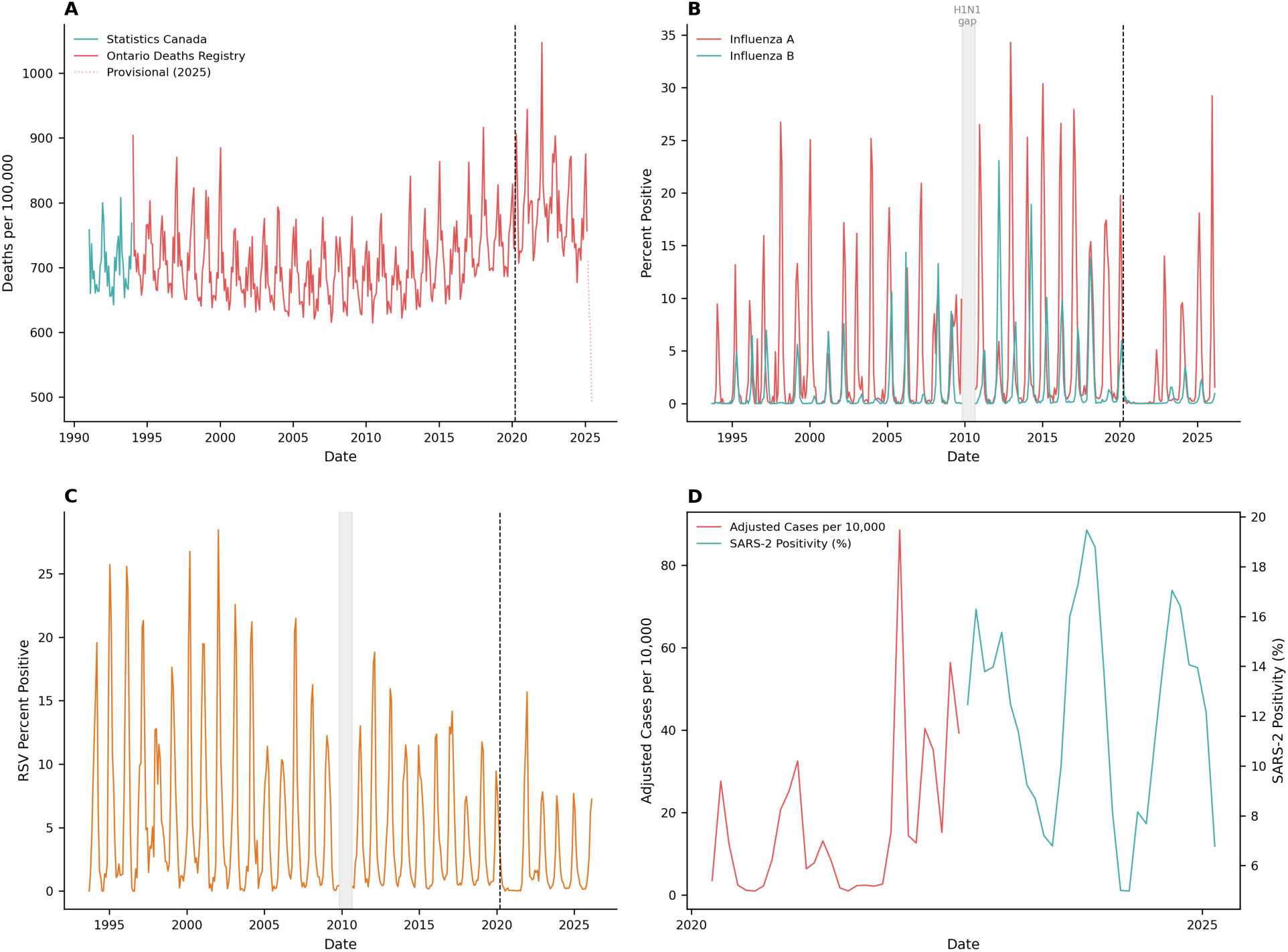
Mortality and virological surveillance data for Ontario, 1993-2025. Monthly deaths per 100,000 from Ontario Deaths Registry (red) and Statistics Canada (teal), demonstrating close concordance between data sources (mean difference 0.3%). (B) Monthly percent positivity for influenza A (red) and influenza B (teal) from FluWatch surveillance, showing regular seasonal epidemics and marked suppression during 2020-2021. (C) Monthly RSV percent positivity from FluWatch surveillance, demonstrating seasonal patterns and pandemic-era disruption. (D) SARS-CoV-2 adjusted case counts per 10,000 population (red, left axis) and SARS-2 positivity percentage (teal, right axis) from March 2020 onward, showing multiple pandemic waves. Vertical dashed lines indicate emergence of the SARS-CoV-2 pandemic in Ontario in March 2020. Panels B and C exclude a period from 2009-2010 during the influenza A H1N1 pandemic for which data were not available (gray rectangle). Data truncated at February 2025 to account for reporting delays in mortality time series.

### Primary Model Results

Separate negative binomial models estimated mortality attributable to respiratory viruses in pre-pandemic and pandemic periods (**Table 1**). In pre-pandemic models (Table 1), only influenza A predicted mortality (IRR 1.0037, 95% CI 1.0028-1.0046). Influenza B did not predict mortality, and RSV showed a non-significant inverse association (IRR 0.9992, 95% CI 0.9979-1.0005, P=0.238). During the combined pandemic period (March 2020–February 2025), only SARS-CoV-2 activity predicted mortality. Both SARS-CoV-2 percent positivity (IRR 1.0073, 95% CI 1.0039-1.0106) and adjusted case counts (IRR 1.0029 per 10,000 cases, 95% CI 1.0022-1.0037) predicted mortality. Influenza viruses and RSV did not show significant associations with mortality in the pandemic-period models.

**Table 1.** Mortality Incidence Rate Ratios for Respiratory Viruses Derived from Negative Binomial Models.

| Period | Virus | Mortality Incidence<br>Rate Ratio (IRR)* | IRR 95%<br>Confidence<br>Interval | Population<br>Attributable<br>Fraction<br>(PAF) | PAF 95% CI | P-value for<br>Association<br>† |
| --- | --- | --- | --- | --- | --- | --- |
| **Pre-pandemic**<br>**(1993 to**<br>**February 2020)** | Influenza A | 1.0037 | 1.0028–1.0046 | 1.821 | 1.389 to 2.251 | <0.001 |
|  | Influenza B | 1.0009 | 0.9992–1.0026 | 0.153 | -0.135-0.441 | 0.297 |
|  | RSV | 0.9992 | 0.9979–1.0005 | -0.453 | -1.210-0.299 | 0.238 |
|  | Combined | --- | --- | 1.531 | 0.455-2.295 | 0.005 |
|  | Respiratory<br>Viruses |  |  |  |  |  |
| **Pandemic** | SARS-CoV- | 1.0029 | 1.0022–1.0037 | ‡6.477 | 4.482-8.429 | <0.001 |
| **(March 2020 to** | 2 (2020- |  |  |  |  |  |
| **February 2025)** | 2022) |  |  |  |  |  |
|  | SARS-CoV- | 1.0073 | 1.0039–1.0106 |  |  | <0.001 |
|  | 2 (2022- |  |  |  |  |  |
|  | 2024) |  |  |  |  |  |
|  | Influenza A | 1.0025 | 0.9985–1.0065 |  |  | 0.219 |
|  | Influenza B | 1.0007 | 0.9789–1.0230 |  |  | 0.95 |
|  | RSV | 1.0032 | 0.9990–1.0073 |  |  | 0.133 |
|  | Combined | --- | --- | 7.612 | 5.145-10.015 | <0.001 |
|  | Respiratory |  |  |  |  |  |
|  | Viruses |  |  |  |  |  |

**NOTE:** \*All incidence rate ratios are per percent increase in test positivity except for SARS-CoV-2 (2020-2022), which is incidence rate ratio per 10,000 increase in adjusted cases.
† P-values for multivariable model coefficients and PAF from PUNAF are identical.
‡ SARS-CoV-2 PAF estimated by setting both early (adjusted cases) and later (percent positivity) model terms to zero.

### Sensitivity Analysis---Models Without Seasonal Adjustment

To evaluate whether seasonal adjustment terms competed with viral predictors for explanatory power, we re-estimated all models excluding Fourier transforms (Table 2). Formal comparison of coefficients using Wald tests revealed substantial differences for viruses with strong seasonal patterns. In the pre-pandemic period, removal of Fourier transforms significantly altered estimates for influenza A and RSV. The influenza A incidence rate ratio increased from 1.0037 to 1.0060 (Wald P<0.001). Most notably, RSV shifted from a counterintuitive non-significant inverse association (IRR 0.9992) to a positive association (IRR 1.0033, 95% CI 1.0023–1.0042, P<0.001), with this reversal being highly significant (Wald P<0.001). Influenza B estimates were unchanged (Wald P=0.497). During the pandemic period, the early SARS-CoV-2 indicator (adjusted cases) increased from 1.0029 to 1.0035 (Wald P=0.042). RSV again demonstrated a significant difference, with the IRR increasing from 1.0032 (P=0.133) to 1.0073 (95% CI 1.0030–1.0116, P=0.001; Wald P=0.006). Other viral predictors did not differ significantly between models.

**Table 2.**
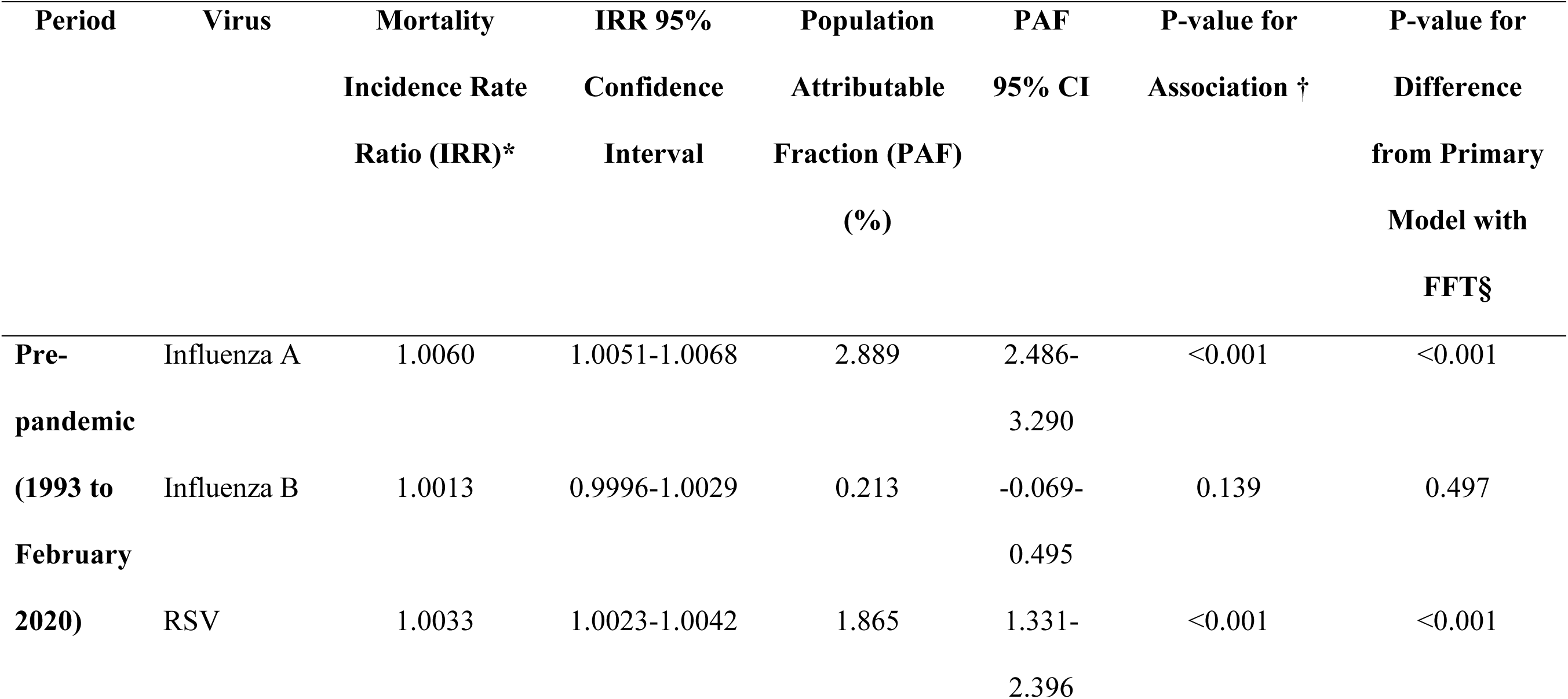
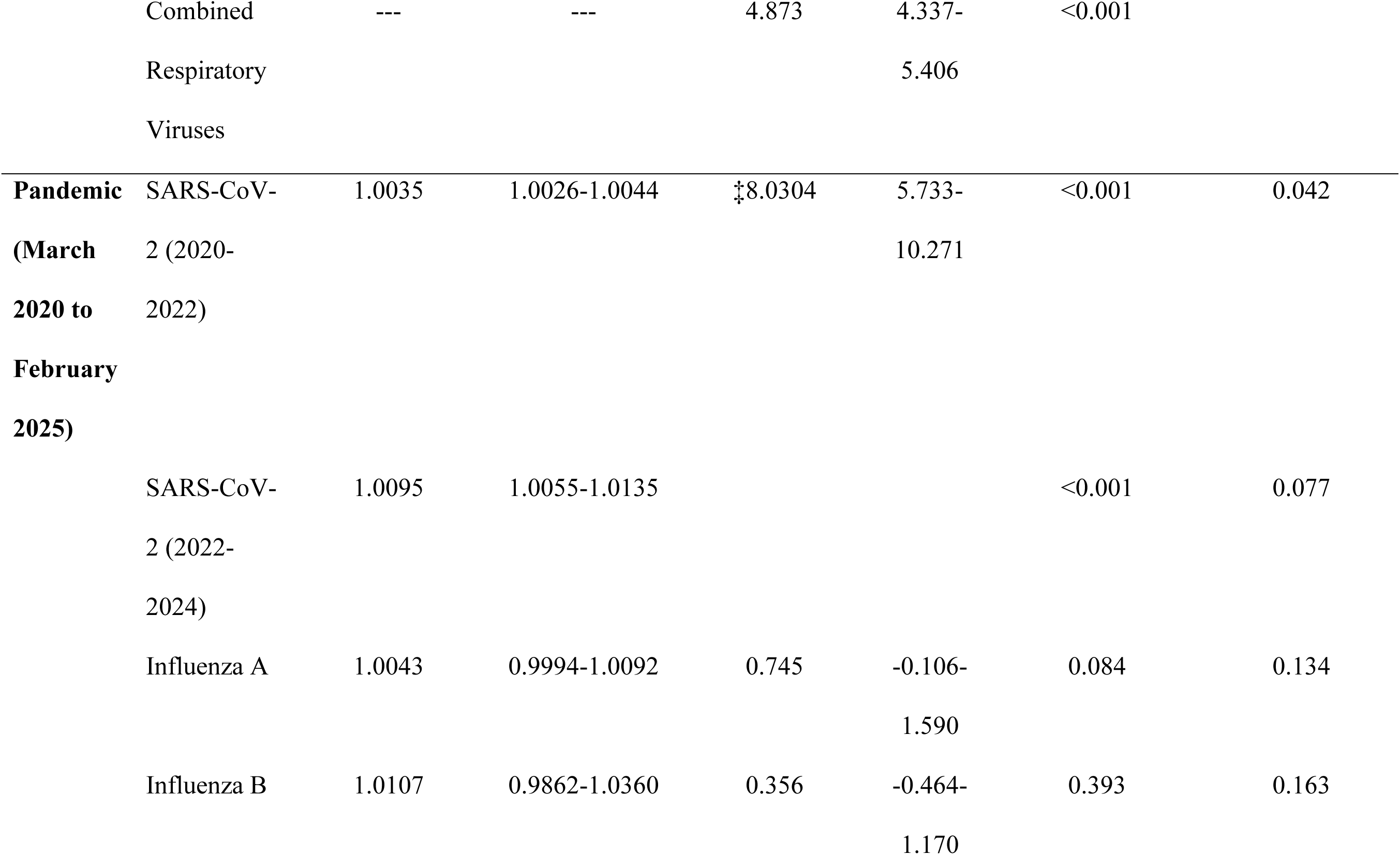
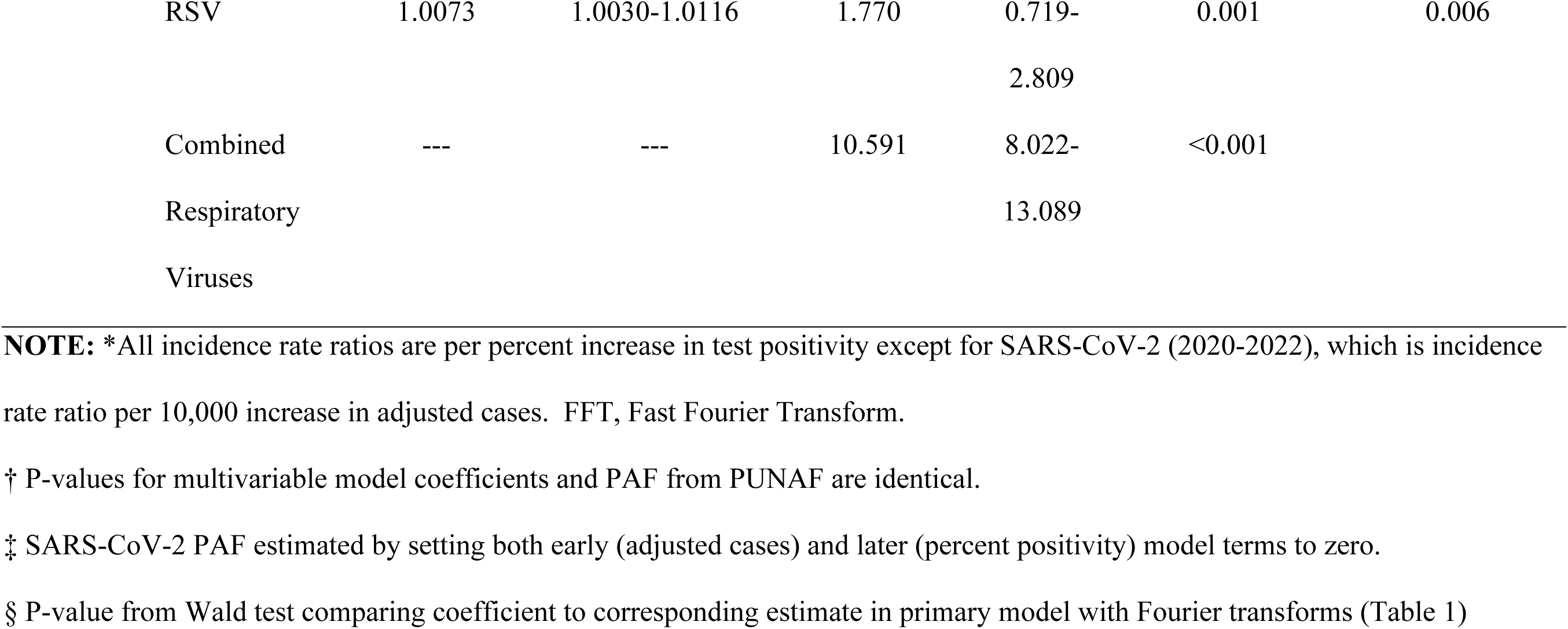
Mortality Incidence Rate Ratios for Respiratory Viruses Derived from Negative Binomial Models without Fast Fourier Transforms.

In a sensitivity analysis using unadjusted SARS-CoV-2 case counts in place of test-adjusted counts, model fit worsened (AIC = 5162.1 vs. 5145.2), supporting the use of test-adjusted data for estimating attributable mortality.

### Population Attributable fraction

Using the primary models with seasonal adjustment (Table 1), we calculated the population attributable fraction for each virus. In the pre-pandemic period, influenza A accounted for 1.8% (95% CI 1.4–2.3%) of all-cause mortality. During the combined pandemic period (March 2020–February 2025), SARS-CoV-2 accounted for 6.1% (95% CI 4.2–8.0%) of all-cause mortality — approximately 4-fold the pre-pandemic influenza A burden. The total number of SARS-CoV-2–attributable deaths predicted by our model closely matched the number of COVID-19 deaths reported by Public Health Ontario over the same interval, providing external validation of our approach. Combined respiratory viruses accounted for 1.5% of mortality pre-pandemic and 7.6% during the combined pandemic period. A sensitivity analysis excluding March–June 2020 yielded a conservative SARS-CoV-2 PAF of 5.7% (95% CI 3.3–8.1%), confirming that the primary estimate is not driven by the extraordinary early pandemic phase. In exploratory temporal stratification, the PHEIC period (March 2020–April 2023; n=38) yielded a SARS-CoV-2 PAF of 7.3% (95% CI 5.0–9.6%) with Fourier adjustment. In the post-PHEIC period (May 2023–February 2025; n=22), SARS-CoV-2 percent positivity remained a significant predictor of mortality (PAF 9.8%, 95% CI 1.1–17.7%; p=0.027), consistent with ongoing endemic SARS-CoV-2 mortality burden. Post-PHEIC influenza A and B attributable fractions did not differ significantly from pre-pandemic baselines (Flu A PAF 1.3%, 95% CI −0.7–3.3%; Flu B PAF 0.6%, 95% CI −1.5–2.7%). Primary inference rests on the combined pandemic model. **Figure 2a** illustrates the temporal patterns of virus-attributable mortality, demonstrating that pre-pandemic combined viral mortality closely tracked influenza A, while pandemic-era combined viral mortality was dominated by SARS-CoV-2.

**Figure 2.**
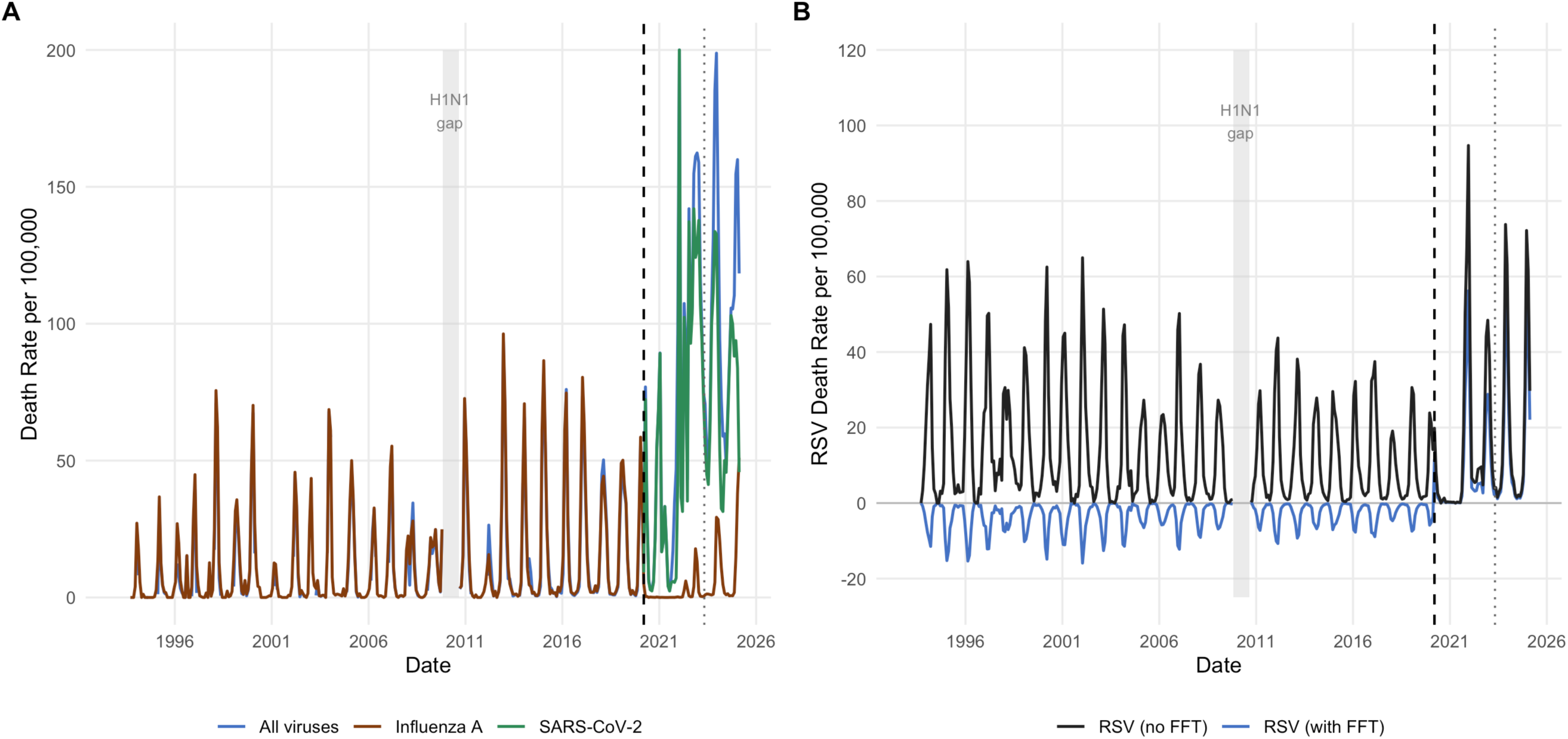
Model-estimated virus-attributable mortality over time. (A) Monthly death rates per 100,000 population attributable to respiratory viruses from primary models with Fourier seasonal adjustment. Blue line shows all respiratory viruses combined, brown line shows influenza A, and green line shows SARS-CoV-2. Pre-pandemic, the combined viral mortality (blue) nearly overlaps with influenza A (brown), demonstrating that influenza A accounted for essentially all virus-attributable mortality. During the pandemic, combined viral mortality (blue) tracks closely with SARS-CoV-2 (green), illustrating SARS-CoV-2’s dominance. (B) RSV-attributable death rates comparing models with Fourier adjustment (blue) versus without (black). With seasonal adjustment, RSV shows frequently negative values pre-pandemic (PAF-0.45%) and substantially attenuated positive associations during the pandemic. Without seasonal adjustment, RSV demonstrates consistently positive mortality associations of similar magnitude (∼1.8-1.9%) in both periods. Both panels exclude a period from 2009-2010 during the influenza A H1N1 pandemic for which data were not available (gray rectangle). Vertical dashed lines indicate emergence of the SARS-CoV-2 pandemic in Ontario in March 2020.

In models without seasonal adjustment (**Table 2**), attributable fractions were substantially higher. Pre-pandemic, influenza A accounted for 2.9% (95% CI 2.5–3.3%) and RSV for 1.9% (95% CI 1.3–2.4%) of all-cause mortality; combined respiratory viruses accounted for 4.9% (95% CI 4.3–5.4%). The contrast between RSV estimates with and without seasonal adjustment is shown in **Figure 2b**. During the combined pandemic period, SARS-CoV-2 accounted for 7.6% (95% CI 5.5–9.7%) and RSV for 2.0% (95% CI 1.1–3.0%) of mortality; combined respiratory viruses accounted for 10.8% (95% CI 8.5–13.1%).

### Meta-Analysis of Population Attributable fraction

Random-effects meta-analyses were conducted to generate pooled estimates stratified by modeling approach (with or without Fourier transforms) and time period (pre-pandemic vs. pandemic) (Figure 3). For all respiratory viruses combined, pooled PAF ranged from 1.5% (95% CI 0.5–2.6%) with seasonal adjustment in the pre-pandemic period to 10.8% (95% CI 8.5–13.1%) without seasonal adjustment during the pandemic, with substantial heterogeneity (I²=95.5%, P<0.001). Influenza A demonstrated high heterogeneity across strata (I²=92.8%, P<0.001). Pooled estimates ranged from 0.6% (95% CI −0.0–1.3%) during the pandemic with seasonal adjustment to 2.9% (95% CI 2.5–3.3%) in the pre-pandemic period without seasonal adjustment. The pooled estimate across all strata was 1.7% (95% CI 0.7–2.6%).

**Figure 3.**
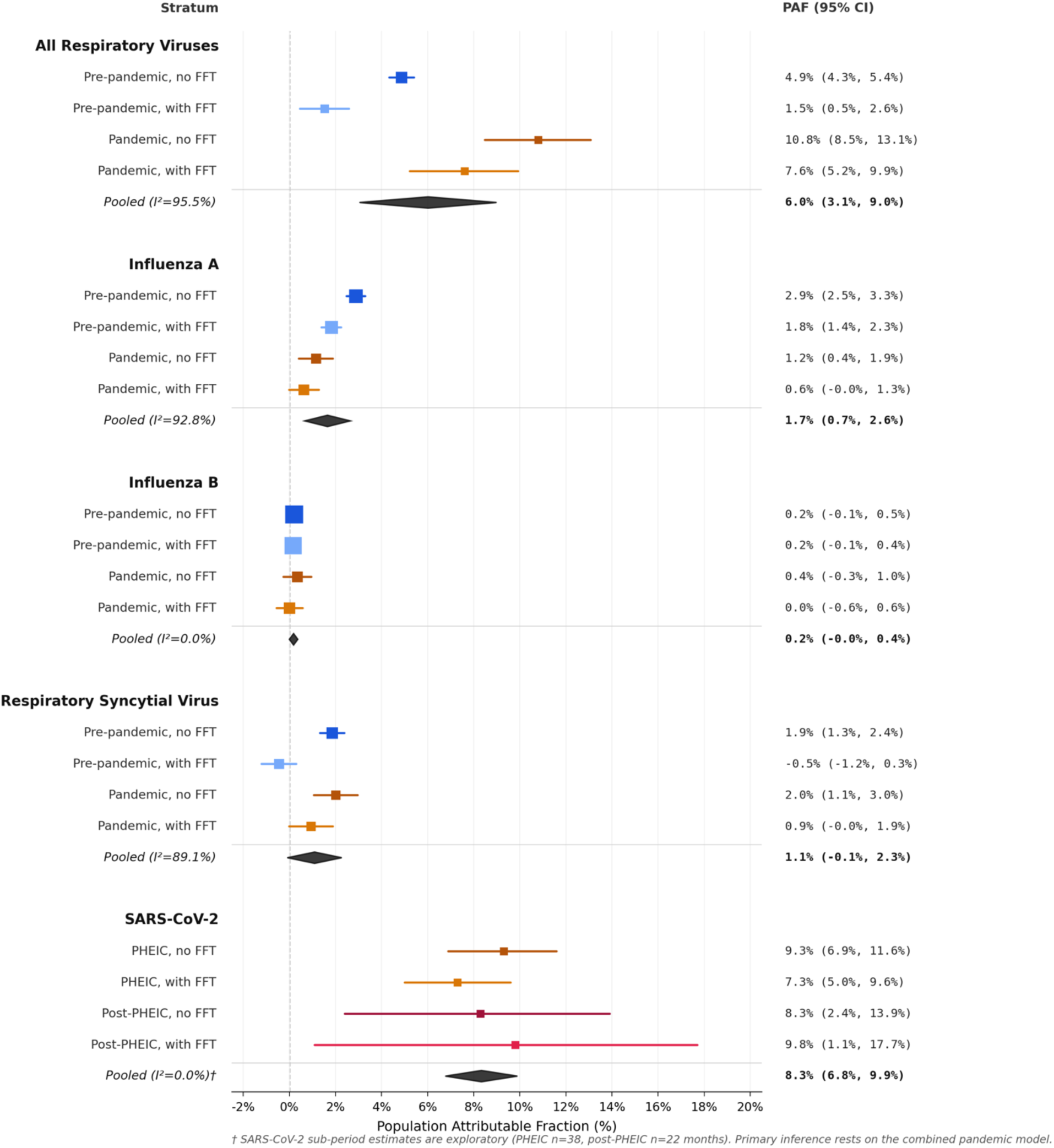
Meta-analysis of population attributable fraction (PAF) for respiratory viruses stratified by modeling approach and time period. Forest plot showing pooled estimates from random-effects meta-analyses for all respiratory viruses combined, influenza A, influenza B, respiratory syncytial virus (RSV), and SARS-CoV-2. For all respiratory viruses, influenza A, influenza B, and RSV, each section includes stratum-specific estimates based on four combinations of modeling approach (with or without Fast Fourier Transform seasonal adjustment) and time period (pre-pandemic: January 1993–February 2020; pandemic: March 2020–February 2025). For SARS-CoV-2, stratum-specific estimates are shown for the PHEIC period (March 2020–April 2023) and post-PHEIC period (May 2023–February 2025), each with and without Fourier adjustment; these sub-period analyses are exploratory given limited sample sizes (n=38 and n=22 months respectively). Diamonds represent point estimates with horizontal lines indicating 95% confidence intervals. Pooled estimates are shown as filled diamonds. I² statistics quantify heterogeneity across strata. FFT, Fast Fourier Transform; PAF, population attributable fraction; PHEIC, Public Health Emergency of International Concern.

Influenza B showed no evidence of mortality association in any stratum, with pooled PAF of 0% (95% CI -0.00–0.00) and no heterogeneity (I²=0.0%, P=0.864). RSV demonstrated substantial heterogeneity (I²=89.1%, P<0.001). Without seasonal adjustment, RSV accounted for approximately 2% of mortality in both pre-pandemic (PAF 0.02, 95% CI 0.01-0.02) and pandemic (PAF 0.02, 95% CI 0.01-0.03) periods. With seasonal adjustment, estimates approached zero or were slightly negative (pre-pandemic: -0.00, 95% CI -0.01-0.00). The pooled estimate across all strata was 1% (95% CI -0.00-0.02). SARS-CoV-2 showed minimal heterogeneity (I²=5.5%, P=0.303) despite different modeling approaches. The pooled PAF was 6.8% (95% CI 5.4–8.3%), ranging from 6.1% with seasonal adjustment to 7.6% without.

## Discussion

In this study, we used more than three decades of mortality and virological surveillance data from Ontario to quantify the proportion of all-cause mortality attributable to major respiratory viruses, extending a previously reported analysis through February 2025. Prior to the emergence of SARS-CoV-2, influenza A was the only virus with a consistent positive association with all-cause mortality after adjusting for seasonal and secular trends, with a PAF of 1.8% (95% CI 1.4–2.3%), consistent with prior studies (16, 17). During the combined pandemic period (March 2020–February 2025), SARS-CoV-2 was associated with a substantially greater population mortality burden, with a PAF of 6.1% (95% CI 4.2–8.0%) in primary models — approximately four times the pre-pandemic influenza A burden — with model-predicted SARS-CoV-2-attributable deaths closely matching reported COVID-19 deaths from Public Health Ontario, supporting the validity of our approach. A sensitivity analysis excluding March–June 2020 yielded a conservative estimate of 5.7% (95% CI 3.3–8.1%), demonstrating that the primary result is not driven by the extraordinary early phase of the pandemic. In contrast, influenza A, influenza B, and RSV did not show significant associations with mortality during the PHEIC period, consistent with the well-documented suppression of seasonal respiratory virus circulation due to SARS-CoV-2 dominance and the implementation of non-pharmaceutical interventions (NPIs) (18, 19).

A key contribution of our study is the formal comparison of models with and without Fourier-based seasonal adjustment. Seasonal terms are commonly used in time-series mortality models to account for periodic fluctuations (10), but because many respiratory viruses also exhibit strong seasonality, these terms may compete with viral predictors for explanatory variance. Similar patterns have been observed in excess mortality studies during the COVID-19 pandemic, where models adjusting for seasonal pathogen circulation produced different estimates than those not accounting for these factors (20). Our findings illustrate this tension: removal of Fourier terms significantly increased estimated effects for pathogens with strong seasonal signatures, particularly influenza A (PAF increased from 1.8% to 2.9%, Wald P < 0.001) and RSV (shifted from PAF –0.45% to PAF 1.9%, Wald P < 0.001).

Meta-analyses revealed substantial heterogeneity for influenza A (I²=92.8%) and RSV (I²=89.1%) across modeling approaches, while SARS-CoV-2 showed minimal heterogeneity (I²=5.5%), indicating a signal robust to modeling choices. Models including Fourier terms provide conservative estimates reflecting mortality variation uniquely attributable to viral indicators after accounting for shared seasonal patterns, while models without such adjustment isolate virus-associated mortality more directly but may conflate viral effects with other seasonal phenomena. The absence of a detectable RSV mortality signal in models with seasonal adjustment initially suggested a statistical artifact driven by viral interference, a well-described phenomenon where staggered epidemic peaks and innate immune responses create negative correlations between viruses (21), but the emergence of positive RSV associations in models without Fourier terms suggests the negative coefficients primarily reflect competition between seasonal and viral covariates rather than true protective effects.

A key new contribution of the extended analysis is characterisation of respiratory virus mortality in the post-PHEIC period (May 2023–February 2025). Point estimates suggested a possible increase in SARS-CoV-2 PAF following formal PHEIC termination, though confidence intervals were wide given the limited sample size (n=22 months), and primary inference is appropriately drawn from the combined pandemic model. Of note, SARS-CoV-2 percent positivity remained a statistically significant predictor of mortality in the post-PHEIC period (PAF 9.8%, 95% CI 1.1–17.7%; p=0.027), suggesting that SARS-CoV-2 continues to exert measurable population mortality burden even after the formal end of the PHEIC. In contrast, post-PHEIC attributable fractions for influenza A (PAF 1.3%, 95% CI −0.7–3.3%) and influenza B (PAF 0.6%, 95% CI −1.5–2.7%) did not differ significantly from pre-pandemic baselines, providing no support for a sustained post-pandemic influenza mortality rebound in this population. Influenza B showed little evidence of measurable mortality impact over the full study period, aligning with prior studies showing it causes milder illness with a smaller population-wide mortality footprint than influenza A (22). The magnitude of SARS-CoV-2’s mortality burden is notable: over the combined pandemic period, SARS-CoV-2 was associated with approximately 4-fold higher attributable mortality than pre-pandemic influenza A, despite facing countermeasures unavailable for historical influenza seasons including non-pharmaceutical interventions (23, 24), multiple vaccine formulations (25), antivirals (26), and substantial partial immunity from prior infection (27). The close concordance between model-based and reported COVID-19 deaths likely reflects the unique features of SARS-CoV-2 surveillance: unlike influenza and RSV, which are under-attributed in routine cause-of-death coding (10, 16), COVID-19 deaths were subject to intensive clinical, laboratory, and public health scrutiny during this period.

Strengths of this analysis include the use of more than 30 years of mortality data, integration of laboratory surveillance indicators, formal evaluation of modeling assumptions through sensitivity analyses and Wald tests, and an extended time horizon now encompassing the post-PHEIC period. Limitations include the use of virological surveillance indicators as imperfect proxies for true infection incidence, the ecological design which cannot establish individual-level causation and assumes viral effects are independent and additive (potentially missing interactions such as interference or co-infections (28)), and the use of all-cause mortality aggregates which mask age-specific patterns and likely underestimate mortality attributable to age-targeted pathogens such as RSV; age-specific mortality attribution will be addressed in future work. The transition in SARS-CoV-2 surveillance from case-based to test-based reporting in September 2022 introduces measurement heterogeneity, which is addressed by including both measures in the pandemic model and setting both to zero in the counterfactual. Statistical power is limited in subperiod analyses given smaller sample sizes. Additionally, mortality data for recent months are subject to reporting delays, which we mitigated by restricting analyses to February 2025.

Despite these limitations, our findings have clear public health relevance. During 2020–2025, SARS-CoV-2 was associated with a population mortality burden 3–4 times higher than pre-pandemic influenza A, a pathogen that itself has historically caused an estimated 3,500 Canadian deaths annually (1). Influenza A remains an important contributor warranting continued seasonal vaccination efforts, while RSV’s consistent ∼2% burden in models without seasonal adjustment and its concentration in vulnerable age groups highlights opportunities for emerging prevention strategies including vaccines for older adults and monoclonal antibodies for infants. The data suggest that SARS-CoV-2 continues to be associated with substantially greater population-level mortality burden than seasonal influenza, even after the formal end of the PHEIC. The persistence of significant SARS-CoV-2-attributable mortality in the post-PHEIC period underscores the need to sustain surveillance and prevention strategies beyond the formal emergency phase. More broadly, this study illustrates the value of long-term respiratory virus surveillance and the importance of modelling choices when quantifying virus-attributable mortality, emphasizing the need for continued investment in surveillance and analytic methods to monitor shifts in viral circulation, quantify preventable mortality burden, and guide immunization strategies.

## References

1. Public Health Agency of Canada. Flu (influenza): For health professionals Government of Canada2024 [updated 2024-10-07. Available from: https://www.canada.ca/en/public-health/services/diseases/flu-influenza/health-professionals.html.

2. World Health Organization. WHO Coronavirus (COVID-19) Dashboard [Available from: https://data.who.int/dashboards/covid19/deaths?n=c&m49=124.

3. Statistics Canada. Deaths, 2022. The Daily2023.

4. Abrams EM, Doyon-Plourde P, Davis P, Brousseau N, Irwin A, Siu W, et al. Burden of disease of respiratory syncytial virus in infants, young children and pregnant women and people. Can Commun Dis Rep. 2024;50(1-2):1–15.

5. ElSherif M, Andrew MK, Ye L, Ambrose A, Boivin G, Bowie W, et al. Leveraging Influenza Virus Surveillance From 2012 to 2015 to Characterize the Burden of Respiratory Syncytial Virus Disease in Canadian Adults ≥50 Years of Age Hospitalized With Acute Respiratory Illness. Open Forum Infect Dis. 2023;10(7):ofad315.

6. Tuite AR, Simmons AE, Rudd M, Cernat A, Gebretekle GB, Yeung MW, et al. Respiratory syncytial virus vaccination strategies for older Canadian adults: a cost–utility analysis. Canadian Medical Association Journal. 2024;196(29):E989–E1005.

7. Mansournia MA, Altman DG. Population attributable fraction. BMJ. 2018;360:k757.

8. Lin CK, Chen ST. Estimation and application of population attributable fraction in ecological studies. Environ Health. 2019;18(1):52.

9. Mieno MN, Tanaka N, Arai T, Kawahara T, Kuchiba A, Ishikawa S, et al. Accuracy of Death Certificates and Assessment of Factors for Misclassification of Underlying Cause of Death. J Epidemiol. 2016;26(4):191–8.

10. Zhou H, Thompson WW, Viboud CG, Ringholz CM, Cheng PY, Steiner C, et al. Hospitalizations associated with influenza and respiratory syncytial virus in the United States, 1993-2008. Clin Infect Dis. 2012;54(10):1427–36.

11. Simcoe Muskoka District Health Unit. Case and Contact Management System (CCM) HealthSTATS SIMCOE MUSKOKA [Available from: https://www.simcoemuskokahealth.org/Health-Stats/HealthStatsHome/Resources/DataSources/CCM.

12. Fisman DN, Greer AL, Brankston G, Hillmer M, O’Brien SF, Drews SJ, et al. COVID-19 Case Age Distribution: Correction for Differential Testing by Age. Ann Intern Med. 2021.

13. Bosco S, Peng A, Tuite AR, Simmons A, Fisman DN. Impact of adjustment for differential testing by age and sex on apparent epidemiology of SARS-CoV-2 infection in Ontario, Canada. BMC Infect Dis. 2025;25(1):589.

14. Statistics Canada. Table 17-10-0009-01: Population estimates, quarterly [Internet]. Government of Canada; Available from: 10.25318/1710000901-eng. Last accessed July 3, 2025. 2025.

15. Government of Ontario. Vital events data by month [Internet]. Ontario Data Catalogue; Available from: https://data.ontario.ca/en/dataset/vital-events-data-by-month/resource/97622ce6-c06a-4970-afe5-be540c748f24. Last accessed July 3, 2025. 2025.

16. Reichert TA, Simonsen L, Sharma A, Pardo SA, Fedson DS, Miller MA. Influenza and the winter increase in mortality in the United States, 1959-1999. Am J Epidemiol. 2004;160(5):492–502.

17. Goldstein E, Viboud C, Charu V, Lipsitch M. Improving the estimation of influenza-related mortality over a seasonal baseline. Epidemiology. 2012;23(6):829–38.

18. Song S, Li Q, Shen L, Sun M, Yang Z, Wang N, et al. From Outbreak to Near Disappearance: How Did Non-pharmaceutical Interventions Against COVID-19 Affect the Transmission of Influenza Virus? Front Public Health. 2022;10:863522.

19. Avolio M, Venturini S, De Rosa R, Crapis M, Basaglia G. Epidemiology of respiratory virus before and during COVID-19 pandemic. Infez Med. 2022;30(1):104–8.

20. Barnard S, Chiavenna C, Fox S, Charlett A, Waller Z, Andrews N, et al. Methods for modelling excess mortality across England during the COVID-19 pandemic. Stat Methods Med Res. 2022;31(9):1790–802.

21. Li K, Thindwa D, Weinberger DM, Pitzer VE. The Role of Viral Interference in Shaping RSV Epidemics Following the 2009 H1N1 Influenza Pandemic. Influenza Other Respir Viruses. 2025;19(4):e70111.

22. Tran D, Vaudry W, Moore D, Bettinger JA, Halperin SA, Scheifele DW, et al. Hospitalization for Influenza A Versus B. Pediatrics. 2016;138(3).

23. Mendez-Brito A, El Bcheraoui C, Pozo-Martin F. Systematic review of empirical studies comparing the effectiveness of non-pharmaceutical interventions against COVID-19. J Infect. 2021;83(3):281–93.

24. Peng A, Bosco S, Simmons AE, Tuite AR, Fisman DN. Impact of community mask mandates on SARS-CoV-2 transmission in Ontario after adjustment for differential testing by age and sex. PNAS Nexus. 2024;3(2):pgae065.

25. Fisman DN, Simmons AE, Tuite AR. Case-cohort design as an efficient approach to evaluating COVID-19 vaccine effectiveness, waning, heterologous immune effect and optimal dosing interval. Vaccine. 2024;42(25):126134.

26. Faust JS, Kumar A, Shah J, Khadke S, Dani SS, Ganatra S, et al. Oral Nirmatrelvir and Ritonavir for Coronavirus Disease 2019 in Vaccinated, Nonhospitalized Adults Aged 18-50 Years. Clin Infect Dis. 2023;77(9):1257–64.

27. Goldberg Y, Mandel M, Bar-On YM, Bodenheimer O, Freedman LS, Ash N, et al. Protection and Waning of Natural and Hybrid Immunity to SARS-CoV-2. N Engl J Med. 2022;386(23):2201–12.

28. Simmons AE, Berry I, Buchan SA, Tuite AR, Fisman DN. Association Between Seasonal Respiratory Virus Activity and Invasive Pneumococcal Disease in Central Ontario, Canada. medRxiv. 2024:2024.09.03.24312990.

